# Reduced survival in COPD patients who received long-term oxygen therapy without meeting prescription criteria: a preliminary study

**DOI:** 10.1101/19000760

**Authors:** François Alexandre, Alain Varray, Yannick Stéphan, Maurice Hayot, Nelly Héraud

## Abstract

**Background:** Randomized clinical trials have provided clear evidence that long-term oxygen therapy (LTOT) increases life expectancy in severely hypoxemic COPD patients. However, in real-life settings, in global COPD cohorts, a paradoxical 2-3-fold increased risk of death has been reported in patients under LTOT. This discrepancy could be explained by a subgroup of patients under LTOT who do not meet the current guidelines and for whom LTOT would be associated with poor prognosis. This retrospective study of a global COPD cohort therefore sought to: (1) determine if a subgroup of patients under LTOT without severe hypoxemia could be distinguished, and (2) compare the mortality risk according to hypoxemia severity and LTOT prescription.

**Method:** The sample was taken from a database (NCT02055885) on 191 stable COPD patients (age: 65.1±9.8 years, 85 women) admitted for a pulmonary rehabilitation program between 2009 and 2012 and followed until 1 January 2018. Uni- and multivariate Cox proportional hazard ratio (HR) models were used to examine the associations between clinical characteristics (age, sex, blood gases, etc.), LTOT according to PaO2 level, and mortality.

**Results:** Forty patients (21%) were under LTOT at PR entry despite not meeting the O2 prescription criteria. Patients under LTOT had a nearly 2-fold higher mortality risk adjusted by covariates (HRmultivariate=1.83; p=0.009). Furthermore, the higher mortality risk under LTOT was specific to the patients under LTOT without severe hypoxemia (HRmultivariate=2.39; p=0.04).

**Conclusion:** The association between LTOT and mortality might be attributed to a subgroup under LTOT despite not/no longer meeting the LTOT criteria. Further studies are needed to identify the physiological basis of this phenomenon.

## Introduction

Long-term oxygen therapy (LTOT) is a proven, well-established therapy to increase life expectancy in chronic obstructive pulmonary disease (COPD) patients with severe hypoxemia. The first data on LTOT’s effects indicated an almost 2-fold increase in survival rates in severely hypoxemic COPD patients undergoing LTOT (PaO_2_≤55 mmHg, or 59 mmHg with concurrent polycythemia or signs of right-sided heart failure or pulmonary hypertension)^1, 2^. While the benefits of O_2_ therapy for severely hypoxemic COPD patients are indisputable, the benefits for the overall COPD population remain unclear. Indeed, a paradoxical association has been reported in global COPD cohorts that include all stages of hypoxemia, with a 2-3-fold increased risk of death in those undergoing LTOT^3-5^. This association remained even after controlling for disease severity (FEV_1_), age, BMI, and exacerbations^3-5^.

Thus, despite LTOT’s substantial clinical impact, its association with a higher risk of death remains unclear as it has not been investigated. Yet if LTOT slows the mortality rate in some types of patients (i.e., severely hypoxemic) but shows an inverse association with the mortality risk in the overall COPD population, this indicates another subgroup of patients who do not meet the current guidelines for LTOT prescription and for whom O_2_ therapy is associated with deleterious consequences. This study of a global COPD cohort therefore sought to: (1) determine whether a subgroup of patients under LTOT without severe hypoxemia could be distinguished, and (2) compare the mortality risk according to the severity of hypoxemia and LTOT prescription.

## Methods

The sample was taken from an unpublished database (NCT02055885) on 191 stable COPD patients (mean age: 65.1±9.8 years, 85 women) who were admitted to the Cliniques du Souffle in France between 2009 and 2012 for a pulmonary rehabilitation program (PR) and followed until 1 January 2018. Blood gases were evaluated on the day patients entered PR, at rest and while breathing room air. No LTOT indications noted by the physician-prescriber were modified, but potential differences with LTOT guidelines were reported by the PR physician in the patient’s medical record. The patients were categorized into 4 groups according to their resting PaO_2_ on air (≤59 mmHg or >59 mmHg, labeled as severe or not-severe hypoxemia: SH or nSH) and LTOT prescription (LTOT or nLTOT):

- nSH-LTOT (PaO_2_>59 mmHg and LTOT)
- nSH-nLTOT (PaO_2_>59 mmHg and no LTOT)
- SH-nTOT (PaO_2_≤59 mmHg and no LTOT)
- SH-LTOT (PaO_2_≤59 mmHg and LTOT)

The baseline clinical characteristics of the 4 groups were compared using ANOVA and HSD post-hoc tests. Time to event was defined as the time (in months) from entry to PR to the month of death or the month when follow-up ended (i.e., 1 January 2018). Univariate and multivariate Cox proportional hazard ratio (HR) models were used to examine the associations between clinical characteristics (age, sex, body mass index: BMI, FEV_1_, FEV_1_/FVC, PaO_2_, PaCO_2_ and 6-minute walking distance: 6MWD), LTOT according to PaO_2_ level, and mortality. Using the means of these covariates in the Cox regression model, the estimated survival functions were calculated for each subgroup. Statistical analyses were conducted with Statistica Software (Statsoft, V13.0, OK, USA). The significance level was set at p≤0.05.

## Results

The median follow-up was 84.7 [Q1: 1.1; Q3: 108.7] months. Descriptive statistics of the population are provided in Table 1. Forty patients were under LTOT (nSH-LTOT group; 21% of the study sample) at PR entry despite not meeting the O_2_ prescription criteria. First, we confirmed that the patients under LTOT had a nearly 2-fold higher mortality risk adjusted by covariates (HR_multivariate_=1.83; 95% confidence intervals: CI=1.16-2.9; p=0.009, data not presented in table). Furthermore, we found that the higher mortality risk under LTOT was specific to the patients under LTOT without severe hypoxemia (nSH-LTOT group; Table 2). In both univariate and multivariate analyses, these patients had an almost 2.5-fold higher mortality risk than the patients without severe hypoxemia and without LTOT (HR_multivariate_=2.39; 95% CI=1.42-4.03; p=0.04). Figure 1 displays the adjusted survival curves for each subgroup.

**Table 1.** Baseline characteristics of the study subjects.

|  | nSH-LTOT (G1) | nSH-nLTOT (G2) | SH-LTOT (G3) | SH-nLTOT (G4) | ANOVA p-value | Significant post-hoc tests (p<0.05) |
| --- | --- | --- | --- | --- | --- | --- |
| n | 40 | 112 | 22 | 17 |  |  |
| Sex (M/F) | 24/16 | 69/43 | 14/8 | 9/8 |  |  |
| Age (years) | 67.2 (10) | 65.7 (10) | 62.1 (8) | 61.7 (8) | 0.11 |  |
| Height (cm) | 166 (10) | 166 (10) | 169 (10) | 165 (8) | 0.52 |  |
| BMI | 25.6 (6) | 25.5 (6.9) | 25.4 (6) | 31.9 (12) | 0.007 | G4 vs G1, G2 & G3 |
| FEV<sub>1</sub> (L) | 0.84 (0.4) | 1.16 (0.5) | 1.04 (0.6) | 1.1 (0.6) | 0.005 | G1 vs G2 |
| FEV<sub>1</sub> (%pred) | 35.3 (15) | 45.7 (19) | 37.4 (24) | 41.6 (20) | 0.02 | G1 vs G2 |
| FEV<sub>1</sub>/FVC | 0.45 (0.16) | 0.5 (0.15) | 0.42 (0.15) | 0.44 (0.16) | 0.07 |  |
| PaO<sub>2</sub> (mmHg) | 69.9 (11) | 68.8 (8) | 53.3 (4) | 55.3 (3) | <0.001 | G2 vs G3 & G4 |
| PaCO<sub>2</sub> (mmHg) | 43.3 (7) | 39.1 (7) | 43.1 (7) | 44.3 (8) | <0.001 | G2 vs G1 & G4 |
| 6MWD (m) | 323 (121) | 397 (116) | 384 (116) | 398 (87) | 0.007 | G1 vs G2 |

BMI: body mass index; FEV<sub>1</sub>: forced expiratory volume in 1 second; FVC: forced vital capacity; 6MWD: 6-minute walking distance; %pred: % of predicted values, nSH-LTOT group: LTOT with resting PaO<sub>2</sub> on air>59 mmHg, nSH-nLTOT group: no LTOT with PaO<sub>2</sub>>59 mmHg, SH-LTOT group: LTOT with PaO<sub>2</sub>≤59 mmHg, SH-nLTOT group: no LTOT with PaO<sub>2</sub>≤59 mmHg. Values are mean ± SD.

**Table 2.**
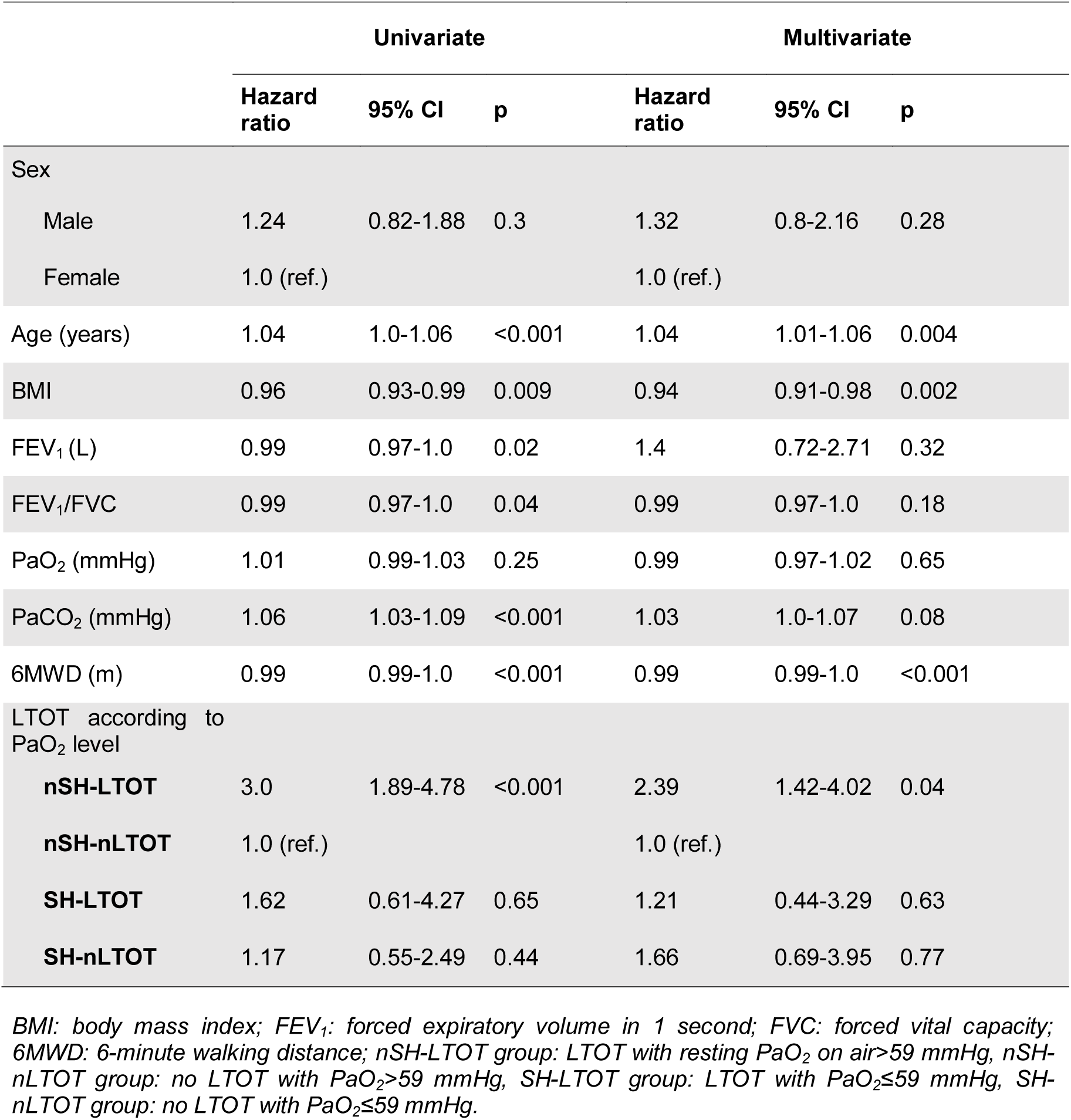
Cox proportional hazard ratio analysis predicting mortality risk.

**Figure 1:**
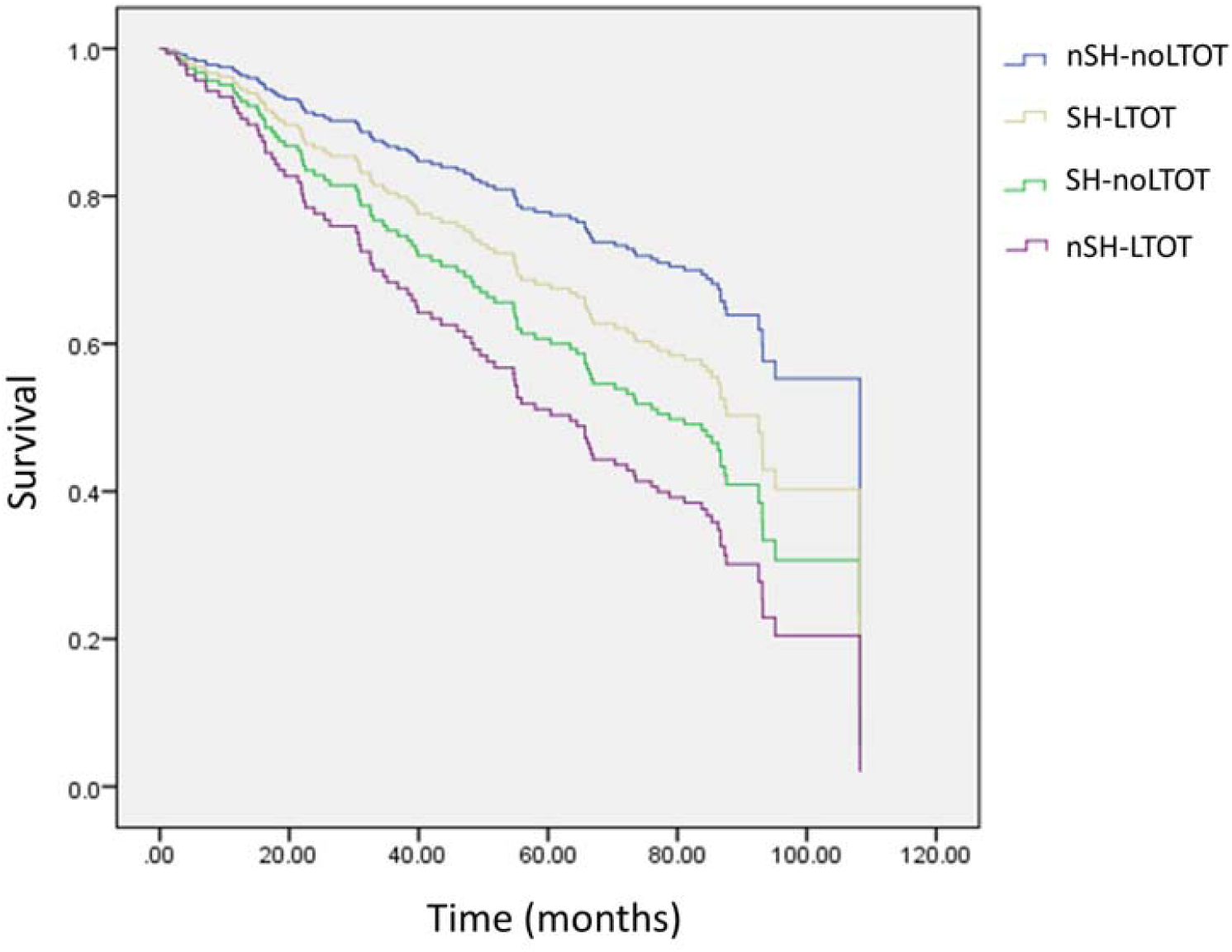
Survival curves in the 4 groups adjusted by age, sex, BMI, FEV1, FEV1/FVC, PaO_2_, PaCO_2_ and 6MWT distance. nSH-LTOT group: LTOT with resting PaO_2_ on air>59 mmHg, nSH-nLTOT group: no LTOT with PaO_2_>59 mmHg, SH-LTOT group: LTOT with PaO_2_≤59 mmHg, SH-nLTOT group: no LTOT with PaO_2_≤59 mmHg.

## Discussion

In line with previous studies, this study showed a higher mortality risk in patients under LTOT in a general COPD population with various hypoxemia severities^3-5^. In addition, we specifically found a nearly 2.5-fold higher mortality risk in the patients under LTOT without severe hypoxemia (PaO_2_>59 mmHg). Therefore, our data shed new light on the paradoxical association between LTOT and mortality in a global COPD cohort, which might be attributed to a subgroup of patients receiving LTOT despite not/no longer meeting the LTOT criteria.

Past research has indicated that COPD patients with PaO_2_>59 mmHg are sometimes prescribed LTOT despite not meeting the indication criteria^6^. This inappropriate prescription has been ascribed to insufficient adherence to guidelines by prescribing physicians^7^. For example, one study found that 18.5% of COPD patients undergoing LTOT had resting PaO_2_ on air >60 mmHg at the time of prescription^8^. LTOT started in unstable patients after hospitalization and poor treatment re-evaluation are also worth considering: ∼40% of patients who meet the hypoxemia criteria at a given time are no longer hypoxemic 1-3 months later^1, 9^. Yet, most patients are never re-evaluated, whether in clinical practice^7^ or longitudinal clinical trials^10^.

This study has limitations. First, the sample size was too small to draw definitive conclusions. Furthermore, comorbidities, adherence to LTOT and exacerbations could not be retrieved. Importantly, although not controlled in this study, the paradoxical association between LTOT and mortality was previously observed regardless of exacerbations^3^.

## Conclusion

This research suggests a plausible explanatory hypothesis about the paradoxical association between mortality risk and LTOT in a general COPD cohort with various hypoxemia severities. In the study, the higher mortality risk under LTOT was specifically found in patients without severe hypoxemia and thus not meeting the current guidelines for LTOT prescription (mean PaO_2_=69.9 mmHg). Larger controlled prospective clinical trials with O_2_ supplementation vs. non-supplementation in the overall COPD population are warranted to identify a physiological explanation for this phenomenon and to rule out (or not) any causal relationship.

## Data Availability

The data are fully available on request to

